# PREACT-digital: Study protocol for a longitudinal, observational multi-center study on wearable- and EMA- based predictors of non-response to CBT for internalizing disorders

**DOI:** 10.1101/2025.03.14.25323957

**Authors:** Leona Hammelrath, Annette Brose, Manuel Heinrich, Pavle Zagorscak, Sebastian Burchert, Till Langhammer, Christine Knaevelsrud

## Abstract

**Introduction:** Despite CBT’s status as a first-line treatment, a substantial proportion of patients does not experience sufficient symptom relief. Recent advances in wearable technology and smartphone integration enable new, ecologically valid approaches to capture dynamic processes in real time. By combining ecological momentary assessment (EMA) with passive sensing of behavioral and physiological information, this project seeks to track daily fluctuations in symptom-associated constructs like affect, emotion regulation and physical activity. Our central goal is to determine whether dynamic, multimodal markers derived from EMA and passive sensing can predict treatment non-response and illuminate key factors that drive or hinder therapeutic change.

**Methods and Analysis:** PREACT-digital is a subproject of the Research Unit FOR 5187 (PREACT), a large multicenter observational study in four outpatient clinics. PREACT channels state of the art machine learning techniques to identify predictors of non-response to cognitive behavioral therapy (CBT) in internalizing disorders. The study is currently running and will end in May 2026. Patients seeking cognitive behavioral therapy at one of four participating outpatient clinics are invited to join PREACT-digital. They can take part in (1) a short version with a 14-day EMA and passive sensing phase prior to therapy, or (2) a long version in which the short version’s assessments are extended throughout the therapy. Participants are provided with a smartwatch and a customized study app. We collect passive data on heart rate, physical activity, sleep, and location patterns. EMA assessments cover affect, emotion regulation strategies, context and therapeutic agency. Primary outcomes on (non)-response are assessed after 20 therapy sessions and therapy end. We employ predictive and exploratory analyses. Predictive analyses focus on classification of non-response, using basic algorithms (i.e. logistic regression, gradient boosting) for straightforward interpretability, and advanced methods (LSTM, DSEM) to capture complex temporal and hierarchical patterns. Exploratory analyses investigate mechanistic links, examine the interplay of variables over time, and analyze change trajectories. Study findings will inform more personalized and ecologically valid approaches to cognitive behavioral therapy for internalizing disorders.

**Ethics and Dissemination:** The study has received ethical approval from the Institutional Ethics Committee of the Department of Psychology at Humboldt Universität zu Berlin (Approval No. 2021-01) and the Ethics Committee of Charité-Universitätsmedizin Berlin (Approval No. EA1/186/22). Results will be disseminated through peer-reviewed journals and presentations at national and international conferences. One year after the last patient out, data will be fully anonymized to allow for open science practices and data sharing, while at the same time securing patients’ data protection rights.

**Trial registration number:** DRKS00030915; OSF PREACT: http://osf.io/bcgax; OSF PREACT-digital: https://osf.io/253nb

**Strengths and limitations of this study:**

1. Large sample of clinical patients starting CBT: Rare in digital phenotyping studies
2. Longitudinal assessment combining passive sensing and EMA: Captures a dynamic, multidimensional view of participants’ behaviors, emotions, and physiological states; Enables analysis of within-person trajectories.
3. State-of-the-art, consumer-grade scanwatches for all participants: Ensures high-quality, standardized data collection across participants; Ensures privacy of participant by not disclosing them as study participants
4. No healthy control group: Limits the ability to distinguish clinical from non-clinical patterns in digital phenotyping data
5. Participant burden due to extensive assessments might decrease adherence and influence therapy outcomes

## INTRODUCTION

The following study protocol describes a subproject from the Research Unit FOR 5187 “Towards Precision Psychotherapy for Non-Respondent Patients: From Signatures to Predictions to Clinical Utility (PREACT)” that was formed as a collaboration across four universities and outpatient clinics in Berlin to identify predictors of non-response to cognitive behavioral therapy (CBT) in Internalizing Disorders. The diagnostic and experimental focus of PREACT lies on emotion regulation as a putative key mechanism of CBT and response to CBT. Due to the comprehensive design of the Research Unit, the general study protocol provides an overview across all subprojects (1). The present protocol entails a more granular description of methods and objectives relevant to this subproject. This level of detail is of utter importance to ensure that reporting recommendations are met (2) and researchers can assess the comparability with other digital phenotyping studies. Associated materials can be found in our OSF directory (https://osf.io/253nb).

Internalizing disorders (ID), such as depressive disorder, obsessive-compulsive disorder, or generalized anxiety disorder, represent the most common mental health condition worldwide (3). Cognitive behavioral therapy (CBT) is considered the first-line treatment for ID, but was shown to produce insufficient response rates (Cuijpers et al., 2024). In addition, urgently needed breakthroughs in psychotherapy research remain elusive: Studies adapting or enhancing CBT or comparing it to other forms of psychotherapy for internalizing disorders often do not find significant differences in outcomes (4).

A great hope lies within the field of machine learning which has found its way into psychotherapy research within recent years (5). Here, a prominent approach is to predict outcomes before the treatment started using a large number of of different variables from various sources (6,7). The idea is to change or adapt a treatment if the risk of non-response is high and thus avoid frustrating experiences for patients as well as unnecessary societal and economical burdens. Thus far, however, results are unsatisfactory: depending on the features of input, predictive accuracies often only slightly exceed chance level, like 62% for neuroimaging markers (8) and/or fail to generalize to external samples (9).

Most existing prediction studies relied on cross-sectional data (i.e., pre-treatment self-reports). However, these momentary snapshots are incapable of depicting the intra-individual symptom-heterogeneity and the inter-individuals therein (10,11). Beyond that, the applicability to outpatient care – as an ultimate goal of precision psychotherapy – is neglected when relying solely on time and cost-intensive forms of phenotyping like clinical interviews or neurophysiological markers (12).

Luckily, recent advances in wearable devices and smartphone technology and the broad integration of smartphones into people’s everyday life are paving the way for more accessible, ecologically valid data collection methods. In particular, these tools offer a scalable alternative that can reduce reliance on labor-intensive procedures. Ecological momentary assessment (EMA) involves short, repeated self-reports on the smartphone. Passive sensing refers to the continuous collection of bio-behavioral data (i.e. physical activity, heart rate) using smartphone or wearable sensors. The combination of active (i.e. EMA-based) and passive (i.e. wearable-based) assessments is also referred to as “Digital phenotyping” (13). Digital Phenotyping offers the opportunity to better understand individual factors and mechanisms leading to (non-) response by (1) opening the black box of between-session processes and gather naturalistic information from everyday settings, (2) getting long-term subjective and objective impressions of symptom dynamics in (3) real time and without recall biases inherent to cross-sectional assessments.

First systematic reviews on digital phenotyping in different internalizing disorders concluded, that, although in its infancy, it can identify behavioral patterns associated with internalizing disorders (14,15). At the same time, only few studies have applied DP in patients undergoing psychotherapy for internalizing disorders to depict (absent) changes in symptoms and/or predict treatment response. De Angel et al. (16) collected DP data in 66 patients starting psychological treatment for depression. They only published results on the feasibility of data collection, showing that data availability varied strongly depending on data source (i.e. smartphone vs. wearable) and treatment stage (i.e. pre-treatment vs. in-treatment vs. post-treatment). Müller-Bardoff et al. (17) conducted a RCT to find sensor– and EMA-based predictors of response to CBT in 150 patients suffering from anxiety disorders. To date, they did not publish results related to DP-based outcome prediction. In psychiatric settings, Zou et al. (18) collected passive sensor data (i.e. phone usage, app usage, sleep) in 245 patients with major depressive disorder to predict response to psychopharmacological outpatient treatment. They were able to achieve sufficient predictive accuracy around 10 weeks before treatment ended.

In summary, there remains a need for studies investigating digital phenotyping (DP) as both predictors and markers of treatment response in internalizing disorders. Our subproject, PREACT-digital, addresses this gap. We implement ecological momentary assessment (EMA) and passive sensing via state-of-the-art wearable devices to continuously monitor patients receiving cognitive behavioral therapy (CBT) for internalizing disorders in their daily lives. Building on the PREACT consortium’s overarching focus on emotion regulation (ER), we incorporate ER-related constructs such as affect and social/situational context into our EMA measures. Ultimately, our goal is to identify ecologically valid, readily implementable markers of treatment response—or the absence thereof—in CBT for internalizing disorders.

Our research objectives encompass predictive and exploratory hypotheses. First, in In line with PREACT, we aim to find out if we can predict NR at T20 and TPost with sufficient predictive accuracy (i.e. >75%) using a combination of EMA and passive sensing features collected during the first, 14-day assessment phase (T0). We expect that EMA-based features can better capture ER-related dynamics and are thus of greatest importance, but passive features will provide a significant incremental value. We further hypothesize, that EMA and sensing features both have an incremental predictive value beyond more elaborate (neuro-)physiological markers and thus represent an ecologically valid substitute for implementation. Second, We aim to find out if symptom changes during CBT can be modeled using EMA and passive sensing, and how they relate to cross-sectional self-report and physiological information. We hypothesize that data actively and passively generated by personal electronic devices such as smartphones and wearables can be linked to neuroimaging-based markers of ER assessed in other subprojects of the research group.

## METHOD AND ANALYSIS

### Sample selection

Participants within the PREACT study are informed about PREACT-digital via flyers and information brochures within each of the participating outpatient clinics. Interested patients are invited to an “onboarding” meeting to review inclusion and exclusion criteria and provide written informed consent. The inclusion criteria of the PREACT study are as follows: (1) Age of 18 years or older, (2) a primary diagnosis based on the DSM-5 criteria for either social anxiety disorder, panic disorder, agoraphobia, generalized anxiety disorder, obsessive-compulsive disorder, post-traumatic stress disorder, or unipolar depressive disorder (major depression or dysthymia), (3) an indication for outpatient CBT treatment, (4) a minimum treatment plan of 12 sessions, (5) a symptom severity indicated by a General Severity Index of the Brief Symptom Inventory (BSI-GSI) greater than 0.56 (19,20), (6) a Clinical Global Impression – Severity Scale (GSI-S) score of 3 or higher, indicating at least mild illness, and (7) owning an appropriate smartphone (i.e. Android version > 9.0 or I-Phone version > 15.0).

Exclusion criteria encompass contraindications for outpatient treatment, including: (1) a current secondary diagnosis of moderate to severe substance use disorder (including regular use of benzodiazepines), (2) current psychosis, (3) current bipolar disorder, (4) more than moderate suicidality, (5) medical conditions that contraindicate CBT. Patients who require inpatient treatment during the course of the outpatient treatment will be excluded.

### Study procedures

Interested patients can choose if they want to take part in PREACT-digital (i.e. therapy-accompanying digital phenotyping). Individuals who decide to participate in the PREACT-digital can choose between two study versions. Option 1 (“the short version”) consists of a 14-day EMA assessment phase paralleled by passive data collection prior to therapy start (T0) only. Option 2 (“the long version”) consists of one additional EMA-phase after 20 therapy sessions (T20) and one after the end of therapy (if the duration of therapy exceeds one year data is collected 365 days after therapy start; TPost). Passive data is collected in parallel throughout the entire time. Participants enrolled in the short version have the option to switch to the long-version after they have completed the first assessment phase. The design of our subproject is depicted in Figure 1.

**Figure 1.**
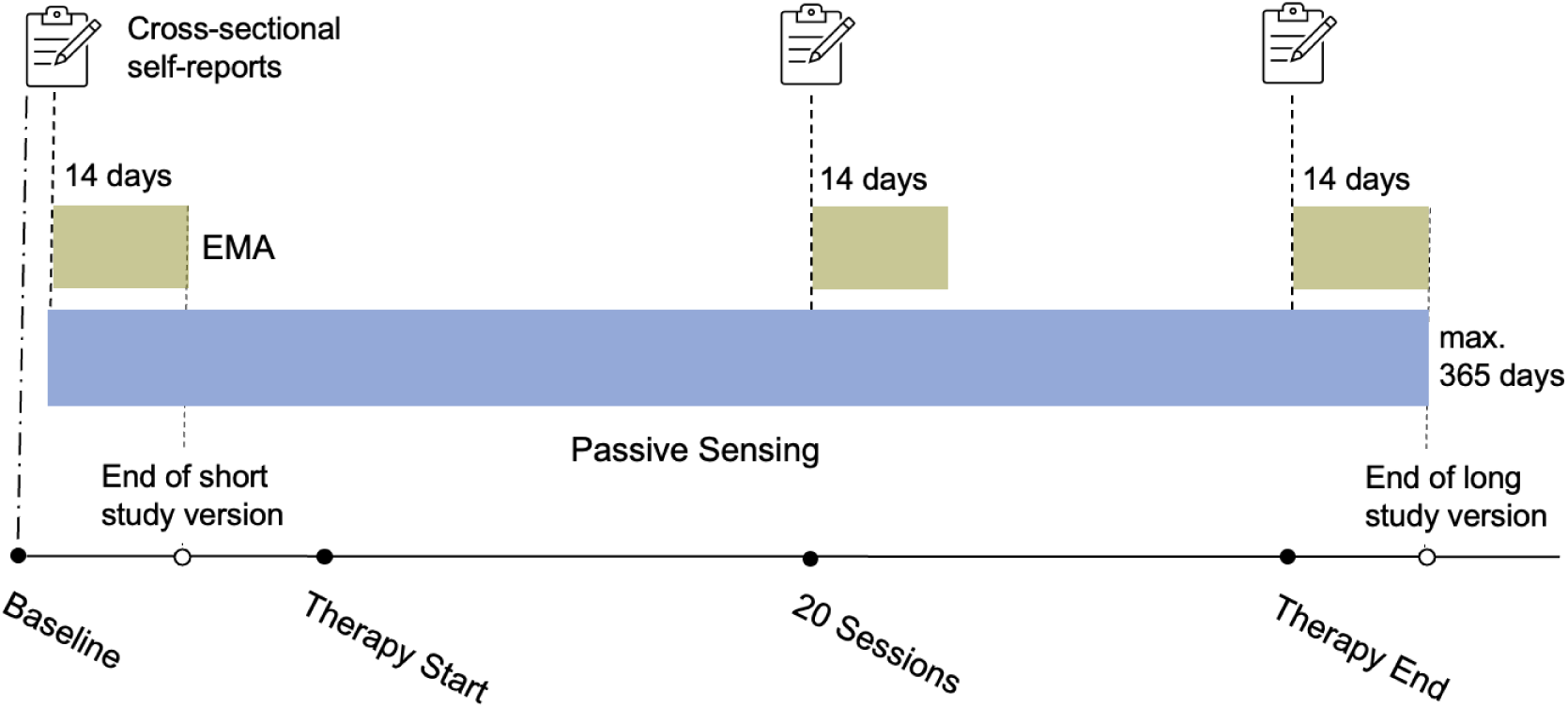
Study Procedures.

Once enrolled in PREACT-digital, participating patients go through an onboarding process with trained research assistants. Together, they install the required apps and connect the smartwatch. For passive-data collection in PREACT-digital, patients receive a state-of-the-art smartwatch (Withings Scanwatch Light), the associated Withings-app and a customized study app developed by a German tech-startup, called TIKI-app. The TIKI-app (1) serves as an interface to the Withings-API, allowing access to the smartwatch data (2) sends out the EMA-questionnaires, and (3) collects GPS data.

Participants receive information and materials for study participation and provide an additional electronic informed consent via the study app. During the first measurement burst, patients receive an onboarding call from the study team, where they get the option to ask questions and report technical problems. In addition, the acceptability and feasibility of EMA and passive sensing is assessed as part of the onboarding call (see interview guideline on https://osf.io/253nb). Information gathered during the onboarding call are entered into a respective REDCap sheet. Between the active assessment phases, patients receive regular update mails with information about their last EMA completion rate and subsequent study procedures.

To reduce interruptions in data transmission leading to high rates of missing data, we implement weekly data monitoring. We run a script to identify participants that have not provided data for more than 7 days. If GPS and/or passive sensing data are absent for more than 7 days, the respective participants receive an e-mail containing instructions on how to restore data transmission. If data transmission is not restored in the following week, participants are contacted by phone. Reasons for interruptions (i.e. app settings, participant forgetting to charge phone) are noted, when available. In addition, all active patients receive a notification on Sunday evenings asking them to open the TIKI-App and check settings to avoid automatic suppression of data collection.

Participants can decide to stop participation in PREACT-digital at any timepoint. In case of dropout, participants are contacted via mail to assess reasons for dropout.

### EMA assessment schedule

During the 14 days of active assessments, patients receive notifications (“beeps”) in the TIKI-App on their smartphones 8-times a day at quasi-random intervals of 90 ± 30 minutes. Depending on their individual sleep-wake rhythm participants can choose to receive beeps between 7.30 and 21.30 or 09.30 and 22.30. The respective questionnaires expire after 30 min. Each EMA assessment contains 30-35 items in total, depending on assessment phase and timing. The order of items was pseudo-randomized.

We employ graded reimbursement, as it was shown to increase compliance: participants received at least 20€ per assessment phase, 35€ if they completed at least 90 beeps, and 50€ if they completed more than 100 beeps. Thus, participants receive up to 150€ for the EMA assessments, which are paid out as a gift voucher. Moreover, individuals participating in the long version of the study are allowed to keep the smartwatch.

### EMA Measures

Table 1 contains an overview of all measures. The whole set of items, including response options and assessment schedules is provided online. Items are presented in German but example items were translated to English here to facilitate comprehension for non-German-speaking readers.

**Table 1:** Overview of collected data.

| Domain | Construct | Description | Example Feature | N Items | Timing, Sampling |
| --- | --- | --- | --- | --- | --- |
| **EMA** | **Positive and Negative Affect** | PANAS-X + item on loneliness | How anxious do you feel right now? | 17 | Baseline, T20, TPost, all beeps |
|  | **Emotion Regulation** | RESS-EMA scale + item on acceptance | In reaction to the negative feeling I...tried to breathe deeply | 8 | Baseline, T20, TPost, all beeps |
|  | **Situational Context** | Self-constructed | How did you spend the time since the last beep? | 1 | Baseline, T20, TPost, all beeps |
|  | **Social Context** | Self-constructed | Did you have social contact since the last beep? | 3 | Baseline, T20, TPost, all beeps |
|  | **Sign. Events** | Self-constructed | How did you perceive the most significant moment since the last beep? | 1 | Baseline, T20, TPost, all beeps |
|  | **Physical Health** | Self-constructed | How physically healthy did you feel today? | 1 | Baseline, T20, TPost, last beep |
|  | **Therapeutic Agency** | Constructed based on TAI | Today I have... implemented ideas or tasks from therapy | 4 | T20, TPost, last beep |
|  | **ECG Control** | Self-constructed | Did you consume coffee in the last 30min ? | 1 | Baseline, T20, TPost, 3rd and 5th beep |
| **Sensing** | **GPS** | Raw; TIKI-App | Time spent at home |  | Continuous; event-based |
|  | **Steps** | Interval sum; Scanwatch | N Steps |  | Continuous; event-based |
|  | **Activity** | Scanwatch | Time spent cycling, calories burned |  | Continuous; event-based |
|  | **Heart Rate** | PPG; Interval mean; Scanwatch | Average heartrate |  | Continuous; regular 30-sec samples |
|  | **ECG** | Resting-State ECG. Scanwatch | Heart Rate Variability |  | Baseline, T20, TPost; 2x daily; 30-sec |
|  | **Sleep** | Aggregate scores; Scanwatch | Time in Bed |  | Continuous |

The following constructs are assessed:

● *Affect*: Participants were asked about their current affective state. The 17 items contain the PANAS-X (21) subscales, with additional single items on loneliness, fatigue and shyness. At each prompt, they were asked to rate on a scale from 1 to 7 how much they currently felt the respective affect.
● *Emotion Regulation*: At each beep, participants were asked to rate the intensity and controllability of their most negative thought since the last beep. Then we assessed the use of different ER strategies since the last beep, using a German translation of the RESS-EMA scale (22), with 6 items covering reappraisal, rumination, suppression, distraction, relaxation. We implemented an additional item on acceptance.
● *Situational Context:* Self-constructed. To assess the situational context, we developed a custom measurement. Participants were asked to specify activities they had pursued in the preceding two hours from a given set of 9 common activities. Participants were able to select multiple options simultaneously. The options were influenced by the DIAMONS scale and a comparable long-term digital phenotyping study (23,24). Here, we aimed to find a balance between sparsity of items and high degree of situational coverage. Subsequently, they were asked to evaluate how much they enjoyed the respective activities.
● *Social Context:* Self-constructed. Participants were asked if they had social contacts since the last beep, how (online/ in person/ phone) and how agreeable the contact was.
● *Significant Events:* Self constructed. Patients were asked to think about the most important event since the last beep and how pleasant they perceived it.
● *Physical fitness:* Self constructed. Participants were asked how physically healthy they had felt today on the last beep of the day.
● *Therapeutic Agency*: We constructed 4 items based on the Therapeutic Agency Inventory (TAI; 25) to assess Therapeutic Agency (TA) in everyday life. The original TAI contains 3 subscales, covering in-session activities, passivity towards the therapist and out-of-session activities. As we were interested in assessing therapeutic agency in everyday life, our TAI-EMA items are based on the “out-of-session activities” subscales and cover cognitive and behavioral aspects of TA.
● *ECG Control:* During measurement bursts, patients were asked twice per day to conduct a resting-state ECG on their Scanwatch. To control for potential confounders influencing the signal, we asked if they had consumed nicotine, caffeine or alcohol or had a heavy meal in the last 30 minutes.

### Passive sensing data

We use a Withings Scanwatch to collect passive data on sleep, physical activity and heart rate (see Table 1 for an overview). The following sensor data are available: for sleep, we have SleepDeepBinary, SleepLightBinary, SleepREMBinary, SleepStateBinary, SleepBinary, SleepInBedBinary, SleepAwakeBinary. They indicate if a user was, for a given time period, in deep sleep, light sleep, REM sleep, any sleep state, asleep overall, in bed (whether asleep or awake), or awake. For physical activity we have ActivityType, ActiveBinary, RunBinary, BikeBinary, WalkBinary, FloorsClimbed, ElevationGain, ActiveBurnedCalories and Steps. ActivityType contains information on a detected activity in a given time period (i.e. resting, walking). The remaining features indicate if, for a given time period, a person was generally physically active, running, biking, walking, how many floors or meters in altitude a person made, how many calories a person burned and the number of steps. Heart rate is collected passively using photoplethysmography (PPG). When no physical activity is detected, the Scanwatch collects heart rate data every 10 min for 30 seconds and provides the average heart rate for that sampling interval. As soon as physical activity is detected, the Scanwatch collects PPG-based heart rate continuously until the activity stops.

The Withings Scanwatch also allows to perform 30-second resting-state ECGs (300Hz) with medical-device quality. During active assessment phases, patients are asked to conduct an ECG on their Scanwatch two times a day. The resting-state ECG data are available in raw format. Withings also calculates Rmssd for resting-state ECGs as an indicator of heart rate variability.

The TIKI-app collects GPS-data directly from the mobile-phone’s sensor. To increase patient’s privacy and data security, GPS data are anonymized locally before saving them on the study servers by rotating the Latitude-Longitude tuples by a random angle. Like this, only mobility patterns (i.e. distance travelled) but not exact locations can be inferred.

### Sample size calculation

As described in the main study protocol (1), the required sample size for the PREACT study was estimated (a) using simulation studies with 5-fold cross-validation to yield a prediction accuracy of 75-80% and (b) using effect size calculation for group-based linear multiple regression analyses, resulting in a required sample of 585 patients. Of those, it was expected that around 80% would take part in PREACT-digital, resulting in an estimated sample size of 468 patients for T0. Based on previous studies investigating the feasibility of digital phenotyping in clinical samples (26), it was further expected that 75% of the sample would decide for the long version of the study, resulting in 350 patients providing data at T20 and TPost.

### Outcomes

We use different machine learning techniques to predict treatment response based on DP data. To evaluate treatment response, we implement two different criteria. First, we follow the definition of the PREACT research group to achieve comparability with other subprojects. It follows the concept of clinically significant change as introduced by Jacobson & Truax (27). The shift from a dysfunctional to a functional mental condition is operationalized by achieving a Brief Symptom Inventory-General Severity Index (BSI-GSI) score of less than 0.56 at T20/post-treatment assessment. Additionally, the change observed from baseline to T20/post-treatment must be reliable, as defined by the Reliable Change Index (RCI). The BSI-GSI is a self-report measure assessed cross-sectionally at T20 and TPost as part of PREACT.

Second, we will use aggregates of EMA assessments on positive and negative affect (PANAS-X; 21) collected during measurement bursts at T20 and TPost as additional indicators of treatment response, as EMA-based measures were shown to capture different aspects of symptom load (28).

To account for the complexity of internalizing disorders, PREACT further included disorder-specific measures as secondary outcomes: The Hamilton Anxiety Rating Scale (HAM-A; 29), the Montgomery–Åsberg Depression Rating Scale (MADRS; 30), the Yale-Brown Obsessive Compulsive Scale (Y-BOCS; 31,32) and the Clinician-Administered PTSD Scale for DSM-5 (CAPS-5; 33,34).

Finally, for exploratory analyses, outcomes will depend on the respective research question, i.e. single-beep scores of EMA constructs or cross-sectional self-reports of emotion regulation capacities.

### Data Preprocessing and Feature Engineering

In its raw format, passive sensing data are usually not interpretable. Thus, depending on the data source, data format and research question, different data preprocessing and aggregation steps will be necessary.

(1) GPS data are collected as tuples containing coordinates (i.e. Latitude and Longitude values). To clean and preprocess the data and build ML features, we follow guidelines by Müller et al. (35). We adapt them to our event-based sampling format, where GPS was assessed when a location-change was detected. From GPS data, we will calculate movement patterns in predefined timescales, i.e. location entropy, distance travelled or time spent at home.
(2) Passive data derived from the Withings Scanwatch will be further preprocessed where possible and necessary, as Withings does not provide raw data for most data sources (see Table 1). Here, we align our preprocessing steps with guidelines for preprocessing sensor data (36). From continuous PPG-based heart-rate data we are able to build features like average HR, min/max HR and HR-zones. From the resting-state ECG we can create more fine-grained features like heart rate variability (HRV). For sleep, we will calculate features describing sleep quality, i.e. sleep duration, proportion of sleep phases and sleep efficiency (i.e. sleep duration/ time in bed) per night. Activity features describe the amount of, or time in active or inactive states, i.e. total number of steps or time spent walking/running/cycling and the amount of calories burned.
(3) For EMA data, we will calculate the Root Mean Square of Successive Differences (RMSSD; i.e. for emotional inertia or emotion regulation variability), individual standard deviations, and mean values. Further feature engineering steps for EMA are described in the *Analysis* section.

Depending on the research question and granularity of data aggregation, a certain compliance rate has to be met to be included in the analysis (i.e. at least 50% of completed EMA beeps). Missing data imputation will also depend on the type of data and applied analyses.

### Analysis

The analysis procedure and code will be provided in our OSF directory. Data preparation, exploration and modelling will mainly be done using Python, R in the Jupyter Lab environment, conducted on the computing facility of Charité – Universitätsmedizin Berlin. In line with recommendations for ML studies and to avoid data leakage, PREACT creates an a-priori holdout set encompassing 20% of participants per subproject. Separate holdout samples for the short and long study version are sampled. For predictive or other analyses encompassing ML, model pipelines will be trained and optimized on the remaining 80% of patients (training set) and evaluated on the holdout set. Exploratory analyses will be performed entirely on the training set.

#### Predictive analyses

We will implement both basic and advanced models and compare subsets of data and different timeframes to balance interpretability and predictive performance. With a sample size of at least 350 participants, we have sufficient data to explore a range of methods. There are various ways to aggregate and preprocess EMA data to include them as predictors of non-response, ranging from simple aggregates to advanced modelling techniques. We will implement dynamic structural equation models (DSEM), which are the current gold standard for modeling interindividual differences in the variable of interest (i.e. emotion regulation) across time. DSEM does not assume equally spaced measurements and consider the time-lag dependence of dynamic parameters such as auto-correlation coefficients and therefore fits the structure data of EMA data perfectly. The Bayesian approach allows for missing data under the missing at random assumption (e.g., when participants skip measurements). Beyond that, DSEM models are extendable to model change in dynamic parameters across measurement bursts. Further, passive data can be flexibly integrated into DSEM to analyze cross-lagged relationships between EMA (i.e. affect) and passive sensing features (i.e. sleep, physical activity). The model-based framework allows us to derive dynamic parameters (e.g., individual autocorrelations, cross-lagged relationships) that we will use as features for the predictive analyses. For prediction of NR, basic algorithms include logistic regression, support vector machines, random forests, and gradient boosting. They are known for its effectiveness on tabular data, require fewer computational resources and are easier to interpret.

Advanced modeling approaches will be selected to incorporate the hierarchical, sequential and multimodal structure of our data. These include deep learning architectures like LSTM, Transformers or ensemble learning approaches. To explore how the predictive capacities depend on the time lag we compare the accuracies of models using subsets of available data (e.g. first measurement burst vs. second measurement burst) and outcome timepoints (i.e. T5 vs. T20 vs. TPost).

To assess feature importance and increase interpretability of our models, we will implement model-agnostic shapley values (SHAP, SHapley Additive exPlanations). In line with the Research Unit, we apply a consistent validation strategy. We repeatedly split data into train, test and validation sets and calculate average performance metrics and variability indices to evaluate the reliability of predictions. In case of imbalanced labels (i.e. response vs. non-response) we apply stratified splits.

#### Exploratory analyses

Our exploratory analyses encompass statistical and inferential investigations, where models are built to explore selected mechanisms and relationships between different variables, data domains and assessment times. We further aim to validate our EMA-based Therapeutic Agency scale using Multilevel Confirmatory Factor Analysis (MCFA). To explore trajectories of change during therapy, we will make use of clustering techniques (i.e. k-means clustering) or latent class growth analysis (LCGA). Derived clusters or classes may serve as additional features for prediction of treatment NR.

## Data handling and dissemination

In compliance with the stringent requirements of the General Data Protection Regulation in Germany (“Datenschutz-Grundverordnung” – DSGVO), we have set up a data protection concept for prospective clinical studies and signed a Joint Controller Agreement by all participating institutions for data handling. These documents have been approved by all respective data protection officers. Given the sensitivity of these health data, they are subject to strict protection protocols and are not shared with third parties as long as they remain identifiable. To ensure the privacy of the patients, all data are safeguarded by appropriate technical (audit-trails and user rights management in access group) and organizational measures (standard operating procedures and standardized personal training) throughout the study phase. One year after the last patient out, data will be fully anonymized to allow for open science practices and data sharing, while at the same time securing patients’ data protection rights. Summaries of findings will also be shared with participants and the public through institutional websites and, where appropriate, public events.

## DISCUSSION

Within the research endeavour of the PREACT study to finding predictors of non-response to cognitive behavioral therapy for internalizing disorders, PREACT-digital implements therapy-accompanying digital phenotyping. DP complements the other subprojects by contributing dense, behavioral and physiological information from the patients everyday life, providing an ecologically valid and innovative add-on to more traditional sources of data. By integrating EMA with wearable-based passive sensing, we capture detailed, real-time information that go beyond traditional cross-sectional snapshots. Our predictive analyses aim to determine whether advanced but computationally demanding modeling approaches improve accuracy over simpler methods, while exploratory analyses focus on uncovering potential mechanisms and subgroups that may inform tailored interventions.

A key strength of our study lies in its ecological validity: participants provide data in naturalistic settings, resulting in rich, high-frequency observations. We further benefit from a relatively large sample size and the opportunity to capture change processes over multiple time points. The integration of our project into the Research Unit allows us to compare our digital phenotypes with other data modalities and acquire a rich, comprehensive impression of participating patients. Although a few studies have integrated DP into outpatient therapy settings, this combination of different data modalities is unique to our study.

We hope to identify both robust predictors of therapy response and clinically meaningful insights that can guide personalized treatment adaptations. As the field of DP in mental health is in its infacy, our study will contribute relevant evidence regarding its feasibility in psychotherapy contexts and its informational content regarding treatment response. Ultimately, this research has the potential to inform the development of more dynamic, precise, and patient-centered therapy approaches for internalizing disorders.

## Author contributions

Drafting the manuscript: LH, AB. Conception & study design: MH, PZ, LH, AB, CK. Acquisition of data: LH, AB. Critical editing and revision of the manuscript: All authors. All authors have approved the submitted version of this manuscript. All authors have agreed to be personally accountable for the author’s own contributions and to ensure that questions related to the accuracy or integrity of any part of the work, even ones in which the author was not personally involved, are appropriately investigated, resolved, and the resolution documented in the literature.

## Patient consent for publication

Not required

## Patient and public involvement

During the first week of study participation, we conduct a standardized interview with participants where we assess the feasibility and acceptability of our ambulatory assessments. Here, patients are also invited to suggest improvements. In addition, as part of another subproject (see SP5 in Langhammer et al., 2024), patients report, among others, the burden and time required for participating in our subproject. All results regarding feasibility and acceptability will be considered when working on the follow-up proposal to this project. To elaborate strategies for dissemination of study results to participants and wider patient communities, we will conduct a congress open to the public in 2025.

## Data Availability

One year after the last patient out, data will be fully anonymized to allow for open science practices and data sharing, while at the same time securing patients data protection rights.

## Acknowledgements

The present work was derived from the Research Unit FOR5187 („Towards precision psychotherapy for non-respondent patients: from signatures to predictions to clinical utility”, www.forschungsgruppe5187.de) funded by Deutsche Forschungsgemeinschaft (project number 442075332). It is based on data from a naturalistic observational clinical study (clinical trial registration: DRKS00030915). We would like to thank the following individuals for their help:

Ulrike Lueken, Lydia Fehm, Norbert Kathmann, Babette Renneberg, Frank Jacobi, Till Langhammer, Andrea Ertle, Björn Elsner, Anne Trösken, Lars Schulze, Ricarda Evens, Chantal Unterfeld, Lena Fliedner, Alexandra Künstler, Torsten Sauder, Leandra Fien, Paul Eichler, Jonathan Torbecke, Lea Sophie Roediger, Freya Uhrlauf, Vera Yuseva, Isabelle Habedank, Nina Richter, Louise Förster, Sophie Meska, Jana Samland, Caroline Nitz, Helen Mahlke, Christoph Geiger, Jasmin Ghalib, Gesa Bimüller, Belnjamin Gas, Dorothea Neumann, Johanna Suchy

## References

1. Langhammer T, Unterfeld C, Blankenburg F, Erk S, Fehm L, Haynes JD, u. a. Design and methods of the research unit 5187 PREACT (towards precision psychotherapy for non-respondent patients: from signatures to predictions to clinical utility) – a study protocol for a multicentre observational study in outpatient clinics. 1. Februar 2025 [zitiert 27. Februar 2025]; Verfügbar unter: https://bmjopen.bmj.com/content/15/2/e094110

2. Trull TJ, Ebner-Priemer UW. Ambulatory assessment in psychopathology research: A review of recommended reporting guidelines and current practices. J Abnorm Psychol. 2020;129(1):56–63.

3. Global, regional, and national burden of 12 mental disorders in 204 countries and territories, 1990–2019: a systematic analysis for the Global Burden of Disease Study 2019. Lancet Psychiatry. 1. Februar 2022;9(2):137–50.

4. Schaeuffele C, Mutak A, Behr S, Fenski F, Hammelrath L, Puetz M, u. a. Increasing the Effectiveness of Psychotherapy in Routine Care through Transdiagnostic Online Modules? Randomized Controlled Trial Investigating Blended Care [Internet]. OSF; 2024 [zitiert 12. März 2025]. Verfügbar unter: https://osf.io/972z5_v1

5. Vieira S, Liang X, Guiomar R, Mechelli A. Can we predict who will benefit from cognitive-behavioural therapy? A systematic review and meta-analysis of machine learning studies. Clin Psychol Rev. 1. November 2022;97:102193.

6. Hornstein S, Forman-Hoffman V, Nazander A, Ranta K, Hilbert K. Predicting therapy outcome in a digital mental health intervention for depression and anxiety: A machine learning approach. Digit Health. 1. Januar 2021;7:20552076211060659.

7. Hammelrath L, Hilbert K, Heinrich M, Zagorscak P, Knaevelsrud C. Select or adjust? How information from early treatment stages boosts the prediction of non-response in internet-based depression treatment. Psychol Med. Juni 2024;54(8):1641–50.

8. Winter NR, Blanke J, Leenings R, Ernsting J, Fisch L, Sarink K, u. a. A Systematic Evaluation of Machine Learning–Based Biomarkers for Major Depressive Disorder. JAMA Psychiatry. 1. April 2024;81(4):386–95.

9. Chekroud AM, Hawrilenko M, Loho H, Bondar J, Gueorguieva R, Hasan A, u. a. Illusory generalizability of clinical prediction models. Science. 12. Januar 2024;383(6679):164–7.

10. Chen LS, Eaton WW, Gallo JJ, Nestadt G. Understanding the heterogeneity of depression through the triad of symptoms, course and risk factors: a longitudinal, population-based study. J Affect Disord. 1. Juli 2000;59(1):1–11.

11. Nemesure MD, Collins AC, Price GD, Griffin TZ, Pillai A, Nepal S, u. a. Depressive symptoms as a heterogeneous and constantly evolving dynamical system: Idiographic depressive symptom networks of rapid symptom changes among persons with major depressive disorder. J Psychopathol Clin Sci. 2024;133(2):155–66.

12. Koutsouleris N, Hauser TU, Skvortsova V, De Choudhury M. From promise to practice: towards the realisation of AI-informed mental health care. Lancet Digit Health. November 2022;4(11):e829–40.

13. Torous J, Bucci S, Bell IH, Kessing LV, Faurholt-Jepsen M, Whelan P, u. a. The growing field of digital psychiatry: current evidence and the future of apps, social media, chatbots, and virtual reality. World Psychiatry. 2021;20(3):318–35.

14. Zierer C, Behrendt C, Lepach-Engelhardt AC. Digital biomarkers in depression: A systematic review and call for standardization and harmonization of feature engineering. J Affect Disord. Juli 2024;356:438–49.

15. Bufano P, Laurino M, Said S, Tognetti A, Menicucci D. Digital Phenotyping for Monitoring Mental Disorders: Systematic Review. J Med Internet Res. 13. Dezember 2023;25(1):e46778.

16. de Angel V, Lewis S, Munir S, Matcham F, Dobson R, Hotopf M. Using digital health tools for the Remote Assessment of Treatment Prognosis in Depression (RAPID): a study protocol for a feasibility study. BMJ Open. Mai 2022;12(5):e059258.

17. Müller-Bardorff M, Schulz A, Paersch C, Recher D, Schlup B, Seifritz E, u. a. Optimizing Outcomes in Psychotherapy for Anxiety Disorders Using Smartphone-Based and Passive Sensing Features: Protocol for a Randomized Controlled Trial. JMIR Res Protoc. 14. Mai 2024;13(1):e42547.

18. Zou B, Zhang X, Xiao L, Bai R, Li X, Liang H, u. a. Sequence Modeling of Passive Sensing Data for Treatment Response Prediction in Major Depressive Disorder. IEEE Trans Neural Syst Rehabil Eng. 2023;31:1786–95.

19. Derogatis LR. Brief Symptom Inventory [Internet]. 2011 [zitiert 12. März 2025]. Verfügbar unter: https://doi.apa.org/doi/10.1037/t00789-000

20. Hiller W, Schindler A, Andor T, Rist F. Vorschläge zur Evaluation regulärer Psychotherapien an Hochschulambulanzen im Sinne der Phase-IV-Therapieforschung. Z Für Klin Psychol Psychother. Januar 2011;40(1):22–32.

21. Haney AM, Fleming MN, Wycoff AM, Griffin SA, Trull TJ. Measuring affect in daily life: A multilevel psychometric evaluation of the PANAS-X across four ecological momentary assessment samples. Psychol Assess. 16. März 2023;35(6):469.

22. Medland H, De France K, Hollenstein T, Mussoff D, Koval P. Regulating Emotion Systems in Everyday Life: Reliability and Validity of the RESS-EMA Scale. Eur J Psychol Assess. Mai 2020;36(3):437–46.

23. Fried EI, Rieble C, Proppert RKK. Building an early warning system for depression: rationale, objectives, and methods of the WARN-D study [Internet]. PsyArXiv; 2022 Aug [zitiert 17. Januar 2023]. Verfügbar unter: https://osf.io/9qcvs

24. Rauthmann JF, Sherman RA. Ultra-Brief Measures for the Situational Eight DIAMONDS Domains. Eur J Psychol Assess. April 2016;32(2):165–74.

25. Huber J, Nikendei C, Ehrenthal JC, Schauenburg H, Mander J, Dinger U. Therapeutic Agency Inventory: Development and psychometric validation of a patient self-report. Psychother Res. 8. Oktober 2019;29(7):919–34.

26. Matcham F, Leightley D, Siddi S, Lamers F, White KM, Annas P, u. a. Remote Assessment of Disease and Relapse in Major Depressive Disorder (RADAR-MDD): recruitment, retention, and data availability in a longitudinal remote measurement study. BMC Psychiatry. 21. Februar 2022;22(1):136.

27. Jacobson NS, Truax P. Clinical significance: a statistical approach to defining meaningful change in psychotherapy research. 1992;

28. Mestdagh M, Dejonckheere E. Ambulatory assessment in psychopathology research: Current achievements and future ambitions. Curr Opin Psychol. 1. Oktober 2021;41:1–8.

29. Hamilton M. The assessment of anxiety states by rating. Br J Med Psychol. 1959;32:50–5.

30. Montgomery SA, Åsberg M. A New Depression Scale Designed to be Sensitive to Change. Br J Psychiatry. April 1979;134(4):382–9.

31. Storch EA, Rasmussen SA, Price LH, Larson MJ, Murphy TK, Goodman WK. Development and psychometric evaluation of the Yale–Brown Obsessive-Compulsive Scale—Second Edition. Psychol Assess. 2010;22(2):223–32.

32. Mataix-Cols D, de la Cruz LF, Nordsletten AE, Lenhard F, Isomura K, Simpson HB. Towards an international expert consensus for defining treatment response, remission, recovery and relapse in obsessive-compulsive disorder. World Psychiatry. Februar 2016;15(1):80–1.

33. Weathers FW, Bovin MJ, Lee DJ, Sloan DM, Schnurr PP, Kaloupek DG, u. a. The Clinician-Administered PTSD Scale for DSM–5 (CAPS-5): Development and initial psychometric evaluation in military veterans. Psychol Assess. 2018;30(3):383–95.

34. Varker T, Kartal D, Watson L, Freijah I, O’Donnell M, Forbes D, u. a. Defining response and nonresponse to posttraumatic stress disorder treatments: A systematic review. Clin Psychol Sci Pract. 2020;27(4):e12355.

35. Müller S, Bayer J, Ross MQ, Mount J, Stachl C, Harari GM, u. a. Analyzing GPS Data for Psychological Research: A Tutorial [Internet]. PsyArXiv; 2021 Juli [zitiert 25. September 2023]. Verfügbar unter: https://osf.io/3cq8n

36. Burns J, Chen K, Flathers M, Currey D, Macrynikola N, Vaidyam A, u. a. Transforming Digital Phenotyping Raw Data Into Actionable Biomarkers, Quality Metrics, and Data Visualizations Using Cortex Software Package: Tutorial. J Med Internet Res. 23. August 2024;26(1):e58502.

